# Structuring clinical text with AI: old vs. new natural language processing techniques evaluated on eight common cardiovascular diseases

**DOI:** 10.1101/2021.01.27.21250477

**Authors:** Xianghao Zhan, Marie Humbert-Droz, Pritam Mukherjee, Olivier Gevaert

## Abstract

Mining the structured data in electronic health records(EHRs) enables many clinical applications while the information in free-text clinical notes often remains untapped. Free-text notes are unstructured data harder to use in machine learning while structured diagnostic codes can be missing or even erroneous. To improve the quality of diagnostic codes, this work extracts structured diagnostic codes from the unstructured notes concerning cardiovascular diseases. Five old and new word embeddings were used to vectorize over 5 million progress notes from Stanford EHR and logistic regression was used to predict eight ICD-10 codes of common cardiovascular diseases. The models were interpreted by the important words in predictions and analyses of false positive cases. Trained on Stanford notes, the model transferability was tested in the prediction of corresponding ICD-9 codes of the MIMIC-III discharge summaries. The word embeddings and logistic regression showed good performance in the diagnostic code extraction with TF-IDF as the best word embedding model showing AU-ROC ranging from 0.9499 to 0.9915 and AUPRC ranging from 0.2956 to 0.8072. The models also showed transferability when tested on MIMIC-III data set with AUROC ranging from 0.7952 to 0.9790 and AUPRC ranging from 0.2353 to 0.8084. Model interpretability was showed by the important words with clinical meanings matching each disease. This study shows the feasibility to accurately extract structured diagnostic codes, impute missing codes and correct erroneous codes from free-text clinical notes with interpretable models for clinicians, which helps improve the data quality of diagnostic codes for information retrieval and downstream machine-learning applications.

## Introduction

The digitization of hospitals has enabled electronic health records (EHR) to become accessible to researchers for secondary usage that benefits healthcare research [1, 2, 3, 4]. The analyses of electronic health records contributes to a better understanding of the clinical trajectories of patients [5], improved patient stratification and risk evaluation [6, 7]. However, much of the information in the EHR is locked in free text clinical notes [2, 4]. Analyzing these free text clinical notes is challenging [1, 2, 8]. Historically, the information in free-text clinical notes has been extracted mostly manually by clinical experts for archiving, retrieval and analyses and this has been particularly relevant to chronic disease as clinical notes dominate over structured data. More recently, natural language processing (NLP) and machine learning methods have shown great promise to automatically analyze clinical notes [1, 2, 9, 10].

EHR data enable researchers and clinicians to perform information extraction and encode the information for later information retrieval and secondary usage [4]. Based on these clinical notes, ICD-10 codes (i.e. the International Classification of Diseases, Tenth Revision) [11] are used by clinicians to encode diagnoses. Some typical research applications of EHR data has been using these diagnostic codes in downstream tasks, such as automatic information retrieval, risk prediction and the prediction of disease subtypes [1, 2, 9, 10]. As the ICD-10 diagnostic codes form the basis, its quality determines the performance of downstream tasks. Furthermore, EHR data in structured format rather than in free-text format can be more easily used in machine learning applications or combined with other data types.

Yet, diagnostic codes are frequently missing in EHR or the recorded diagnostic codes may be inaccurate. Misclassification and inaccuracy in diagnostic codes have been reported in an increasing number of papers, for instance, in cases related to myocardial infarction and stroke [12, 13]. Mccarthy et al. [12] reported that a substantial percentage of patients who had myocardial injury were miscoded as having type 2 myocardial infarction, which may have serious consequences. Next, Chang et al. [13] found disagreement in stroke coding, which may negatively influences stroke case identification in epidemiological studies and hospital-level quality metrics. Recent studies have focused on the problem of diagnostic code prediction [1, 9]. Although some good results have been shown, many of the previous diagnostic code prediction studies have applied deep-learning methods that make the models hard to interpret [2, 9, 3]. Because ICD-10 codes are usually the start for downstream tasks and clinicians attach great significance to interpretable information extraction systems [4], interpretable models may have certain advantages than less-interpretable models in that they may not only enable accurate ICD-10 code imputation but also enable clinicians to readily understand the models and control the quality of the diagnostic codes with their expertise.

In this study, we propose the use of NLP word vectorization algorithms and logistic regression (LR) to predict eight ICD-10 codes related to common cardiovascular diseases from free-text outpatient progress notes. We compared both interpretable models and less interpretable models with regards to their performances on the ICD-10 code prediction tasks. The proposed models show good classification performance on eight ICD-10 codes on two Stanford cohorts and the models generalized well to the MIMIC-III data set. Additionally, the most interpretable models also showed the best performance on all data sets [14, 15].

## Methods

### Data description

We used outpatient progress notes of 133,644 patients diagnosed with cardiovascular diseases at Stanford Health Care. The patients were partitioned into a training set (60%), validation set (20%) and test set (20%). All notes belonging to the same patient were partitioned into the same data set to avoid information leakage across data sets. The data set included 5,604,539 notes from 31,502 encounters dated from April, 2000 to October, 2016. The data was retrospectively collected and de-identified in accordance with approved IRB guidelines by Stanford University (Protocol: IRB-50033 - Machine Learning of Electronic Medical Records for Precision Medicine). (Fig. 1).

**Figure 1:**
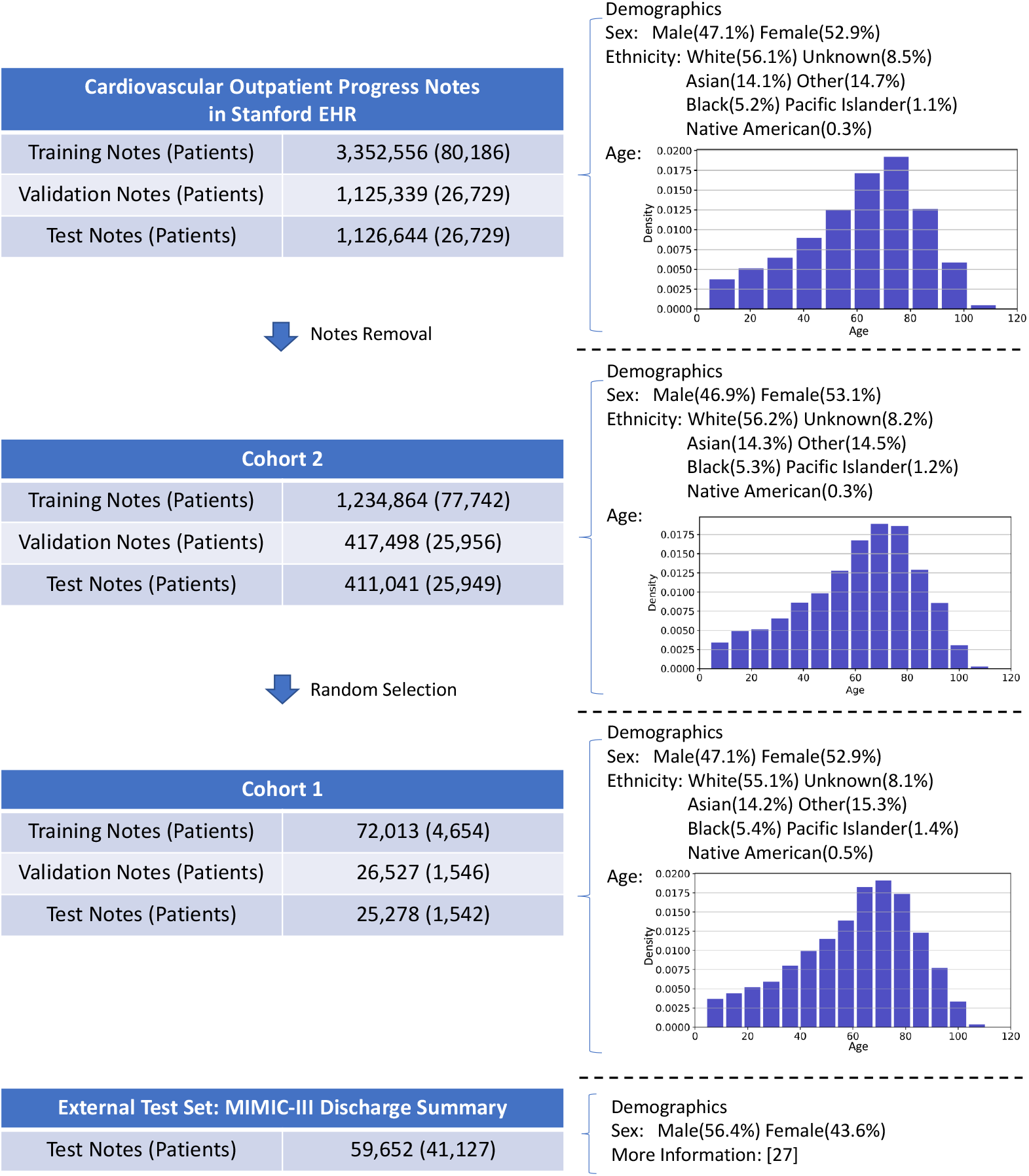
Visualization of the cohorts used in this study with the number of notes and patients of the cardiovascular electronic health records at Stanford, two subsets: Cohort 1 and Cohort 2, and the MIMIC-III test set for model validation. In each cohort, we also show the major demographic features including sex, ethnicity and age.

We focused on the following eight common cardiovascular diseases from clinical notes: acute myocardial infarction (I21), chronic ischemic heart disease (I25), other pulmonary heart disease (I27), cardiomyopathy (I42), atrial fibrillation flutter (I48), heart failure (I50), atherosclerosis (I70), esophageal varices (I85). As ICD-10 codes have a hierarchy to organize the over 69,000 diagnostic codes, we aimed at predicting the three-letter prefixes of the ICD-10 diagnosis codes.

### Data processing

Notes with fewer than sixty words and notes without any labeled ICD-10 code were excluded, resulting in the removal of 63.2% notes defining Cohort 2. For prototyping and testing the scalability of the models, a smaller cohort, Cohort 1 was built with randomly selected notes from Cohort 2 (Fig. 1 and Supplementary Table 1).

Next, we processed the clinical notes by changing them to uncapitalized text and removing any special characters, punctuation, mathematical symbols and universal resource locators (URLs). Stop-words such as conjunctions were removed with Gensim [16] and the words were tokenized. Stemming was then used to reduce inflected words to word stems with Porter stemming algorithm [17] with the NLTK library [18].

### Word embeddings

We used four different vectorization algorithms to convert the free-text notes to numerical features (i.e. word embeddings): Bag-of-words (BOW), term frequency-inverse document frequency (TF-IDF), word2vec (W2V) and doc2vec (D2V). In addition, the batch-word2vec (W2V batch) was introduced as a modified model based on word2vec.

BOW [19] and TF-IDF [14] are word-count based embeddings. In this study, after applying BOW and TF-IDF to Cohort 1 and Cohort 2, the feature dimensions were 88,815 and 414,391 respectively.

W2V [20] is another vectorization algorithm to get word embeddings based on shallow neural networks. In this work, a pre-trained W2V model was used: a biomedical W2V model trained on a corpus collected from PubMed and MIMIC-III [21] with 16,545,452 terms and an embedding dimension of 200. After converting each term in a text into a 200-dimension embedding, an average of all the term embeddings was taken as the embedding for one individual note.

The progress notes we used can be divided in three general sections, describing patient history, description at presentation and plan/billing. In addition to taking the average as a note embedding, a batched form of W2V was introduced in this study by splitting a note into several batches (*n* = 1, 3, 5) to extract section-related contents. For instance, a note with a length of 1,000 words could be split into five batches and the first 200 word embeddings were averaged as the feature of the first batch. In this modified batch-word2vec model (W2V batch), the embedding dimension was 200*n* where *n* was the number of batches. The *n* was chosen to be three based on the average area under receiver operating characteristic curve (AUROC) on the validation set in Cohort 1.

D2V is based on W2V but further inputs the tagged document id in the training of word vectors [22]. In the training process, a word vector is trained for each term, and a document vector is generated for each document. In the inference process for prediction, all weights are fixed to calculate the document vector for a new document. In this study, to avoid overfitting, we used the 63.2% dropped notes (neither in Cohort 1 nor in Cohort 2 because the notes were either shorter than 60 words or without any ICD-10 codes) to train our D2V model with 40 epochs and an embedding dimension of 200. The number of terms modeled was 327,113.

To visualize the data, the nonlinear dimensionality reduction method, t-distributed Stochastic Neighbor Embedding (t-SNE) [23] was used.

### Classification algorithm

Once we get the note embeddings, the vectors become the input of a classification model to predict the diagnostic code. We used logistic regression (LR) for ICD-10 code prediction considering model interpretability. LR [24] applies the logistic function in combination with least square regression for classification. In this study, we used a Python implementation of LR in the scikit-learn package [25]. L2 regularization was used in this study and the penalty strength *C* was tuned based on the average AUROC on the validation set in Cohort 1. A 1:50 class weight was added to deal with the imbalanced cases since the average prevalence of the eight I-codes was approximately 2%.

### Model assessment and interpretation

To assess the performances of different word vectorization methods, AU-ROC and area under precision recall curve (AUPRC) were used as the metrics to evaluate the word embeddings and the LR models in eight diagnostic code classification tasks. On Cohort 1, bootstrapping [26] was done on the training set for thirty times to test the model robustness.

As BOW and TF-IDF are directly interpretable word-based vectorization algorithms, to interpret the models, the LR coefficients were analyzed to identify the important words in classification. The top ten most important words for decision were extracted after bootstrapping the training samples in thirty repeats. In each of the bootstrapping experiments, the thirty most important words were extracted as the candidates, and the final top ten most important words were selected based on two metrics: 1) the ranking metric: the sum of rankings of the important words over all bootstrapping results (smaller ranking sums mean higher importance); 2) the coefficient metric: the sum of LR coefficients of the important words over all bootstrapping results (larger coefficient sums mean higher importance).

Because the recorded diagnostic codes can be missing and inaccurate in clinical practice, to test whether it was possible to impute missing ICD-10 codes based on the model predictions, several false positive cases were randomly selected and the corresponding notes were analyzed.

### External validation

Next, the model transferability was tested on the MIMIC-III (Medical Information Mart for Intensive Care III) data set of de-identified health-related data of 40,000 intensive care unit stays at Beth Israel Deaconess Medical Center [27]. We directly applied the word embedding models (BOW, TF-IDF, W2V, W2V batch and D2V) and the corresponding LR classifiers trained on the training set of the larger Cohort 2 of Stanford notes to predict the diagnostic codes of the discharge summary in MIMIC-III data set (59,652 notes, 41,127 patients). No model fine-tuning on the MIMIC-III data set was done. As MIMIC-III uses the ICD-9 as diagnostic codes, the ground truth was set to the corresponding ICD-9 codes of the eight cardiovascular diseases. In this study, we matched the ICD-10 codes to the corresponding ICD-9 codes by matching the three-letter prefix and the highest hierarchy of the ICD-9 code that describes a specific disease. The matched ICD-9-ICD-10 codes [28] of the diseases and the prevalence in the MIMIC-III discharge summary are: 410 (I21, acute myocardial infarction), 10.36%; 414 (I25, chronic ischemic heart disease), 26.64%; 416 (I27, other pulmonary heart disease), 4.95%; 425 (I42, cardiomyopathy), 3.92%; 427 (I48, atrial fibrillation flutter), 32.16%; 428 (I50, heart failure), 25.98%; 440 (I70, atherosclerosis), 3.61%; 456 (I85, esophageal varices), 1.77%. Proportional z-tests showed statistically significant difference in the prevalence of the eight codes between the Cohort 2 training set of Stanford data and the MIMIC-III data.

## Results

### Data Visualization

We first visualized the feature embeddings with TF-IDF using t-SNE to explore the data in the clinical notes in Cohort 1 training set (Fig. 2). Due to limited space, we presented the TF-IDF visualization as a demonstration because of its full interpretability. We found clusters related to several cardiovascular diseases. The selected clusters within the bounding boxes showed high prevalence in I-codes, suggesting that the feature embeddings may be able to distinguish ICD-10 codes.

**Figure 2:**
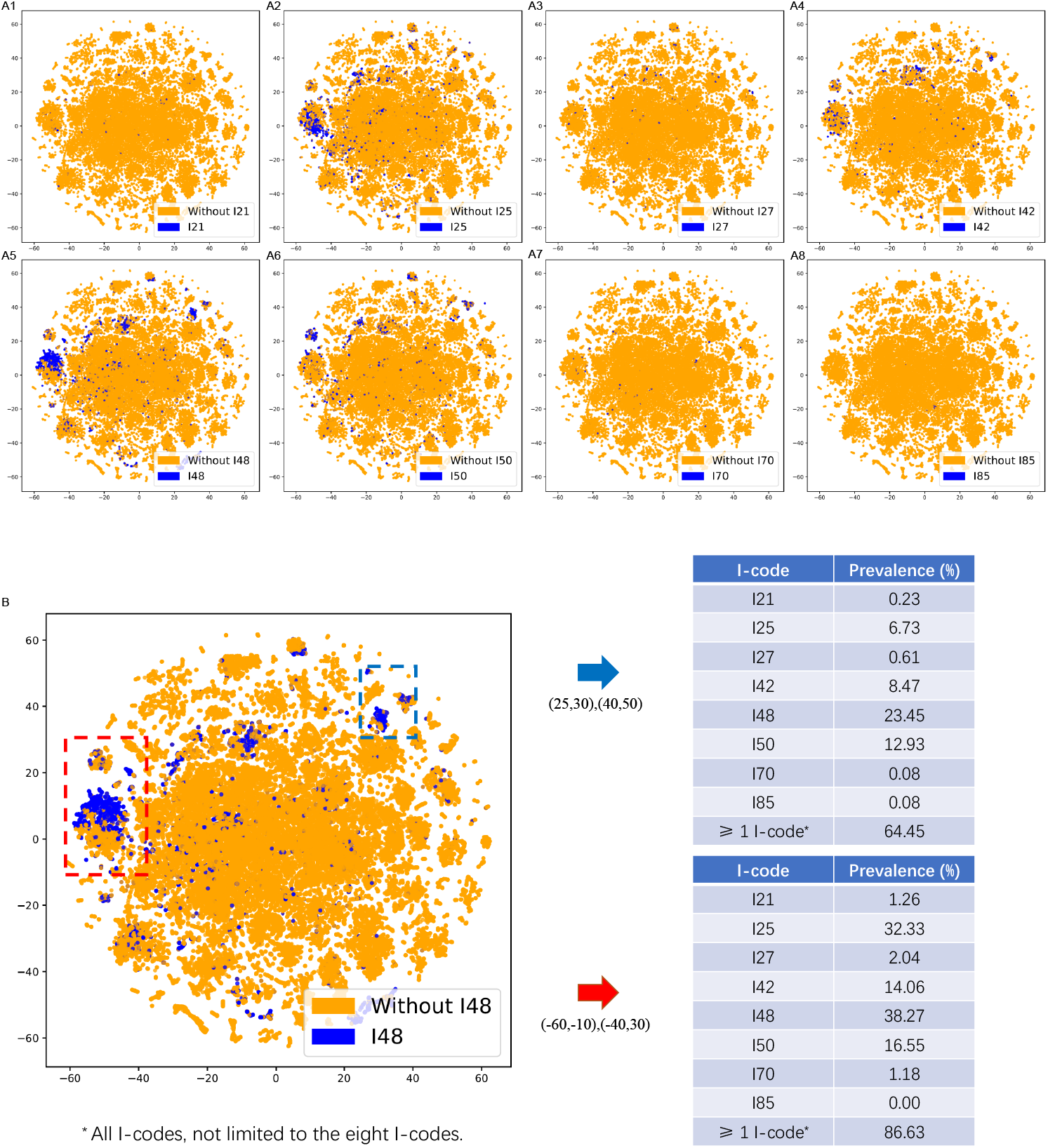
The t-SNE visualization of the training notes in the Cohort 1 of eight I-codes based on TF-IDF. A. The t-SNE visualization of the TF-IDF embeddings of the Cohort 1 training notes of eight I-code classification tasks. B. The prevalence of eight I-codes in the two selected regions with high prevalence of I-codes. Here, ‘more than 1 I-codes’ means all types of I-codes, not limited to the eight I-codes we investigated.

### Prediction of ICD-10 Codes

First, we tested our machine learning workflow for predicting ICD-10 codes on Cohort 1. These results showed that LR and the word embeddings enabled the classification of the eight diagnostic codes related to cardiovascular diseases (I-code) with high prevalence on Cohort 1 with both high AUROC and high AUPRC (Fig. 3, Supplementary Fig. 1). The AUROC values in all classification tasks were higher than 0.75 and TF-IDF outperformed the other four embeddings with AUROC values higher than 0.85 on four selected codes with different prevalence. There was a variance in the AUPRC among the codes with varying prevalence. For the codes with high prevalence such as I25 and I48, the AUPRC values were above 0.60 and 0.70 for TF-IDF. Additionally, the thirty bootstrapping experiments on Cohort 1 showed the best performances given by TF-IDF on the majority of the codes (Fig. 4).

**Figure 3:**
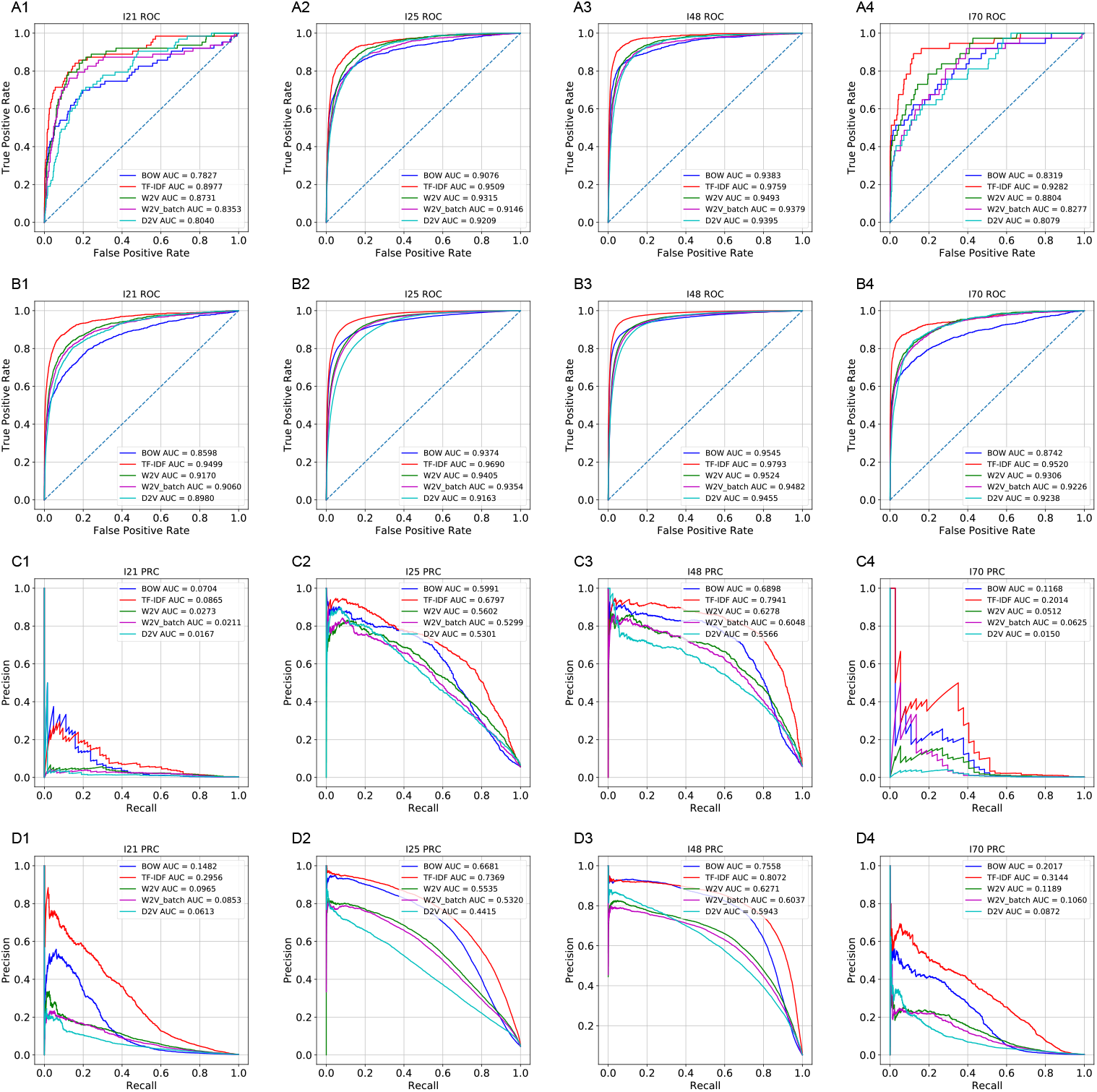
The receiver operating characteristic curves and precision recall curves of the LR models trained on five different word embeddings and on four of the eight I-code classification tasks which represent different prevalence (Cohort 1: A1-A4 and C1-C4; Cohort 2: B1-B4 and D1-D4).

**Figure 4:**
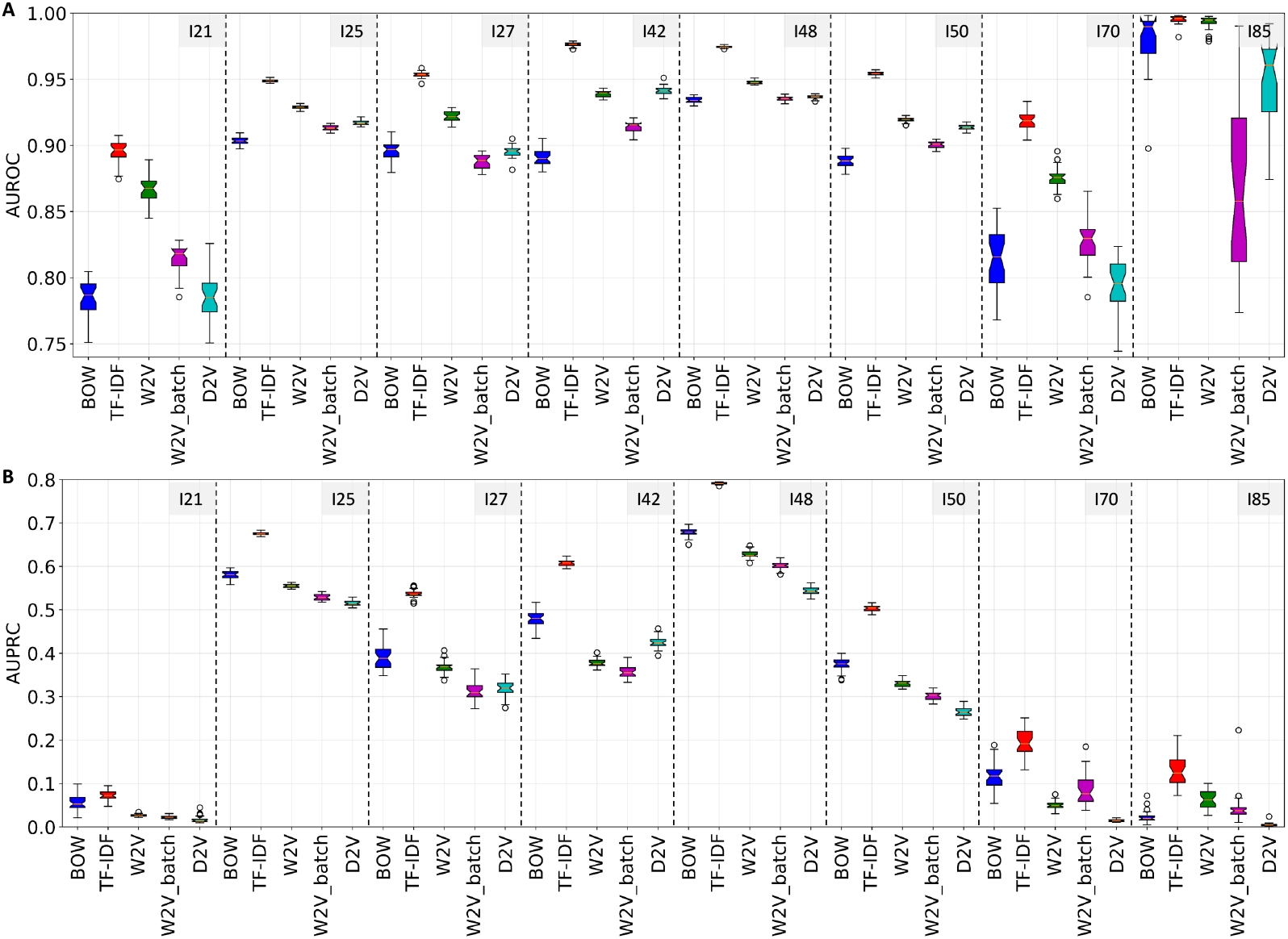
The AUROC and AUPRC of classifiers based on different word embeddings in thirty bootstrapping experiments on Cohort 1. A. The AUROC results: the best model in bootstrapping experiments based on AUROC was TF-IDF (mean AUROC, (95% CI)): I21: 0.8952 (0.8768-0.9075), I25: 0.9487 (0.9470-0.9514), I27: 0.9537 (0.9505-0.9585), I42: 0.9763 (0.9735-0.9790), I48: 0.9745 (0.9731-0.9762), I50: 0.9543 (0.9522-0.9571), I70: 0.9185 (0.9046-0.9333), I85: 0.9951 (0.9918-0.9981). B. The AUPRC results: the best model in bootstrapping experiments based on AUPRC was TF-IDF (mean AUPRC, (95% CI)): I21: 0.0723 (0.0549-0.0951), I25: 0.6752 (0.6709-0.6830), I27: 0.5370 (0.5189-0.5557), I42: 0.6079 (0.5949-0.6240), I48: 0.7913 (0.7878-0.7948), I50: 0.5028 (0.4888-0.5161), I70: 0.1941 (0.1344-0.2514), I85: 0.1281 (0.0727-0.2108).

Secondly, on the larger Cohort 2, with more data, the results showed that the LR models trained on the word vectorization methods classified the I-codes with an improvement in both AUROC and AUPRC, particularly on the codes with lower prevalence (Fig. 3, Supplementary Fig. 2). TF-IDF outperformed the other word embeddings in terms of both AUROC and AUPRC. On the codes with lower prevalence (i.e. I21 and I70) the performances were significantly improved with AUROC values around 0.95 and AUPRC values above 0.25 based on TF-IDF word embeddings.

### Interpretation of important words in classification

To interpret the models, the ten most important words were extracted in thirty bootstrapping experiments on Cohort 1 (Table 1). The results showed that not only many important words that were found were overlapping in the bootstrapping experiments, but also that most words could be explained based on the meanings related to the diagnostic codes. For example, for acute myocardial infarction, non-ST-elevation myocardial infarction, myocardial, myocardial infarction, thrombus and infarction were found important; for chronic ischemic heart disease, coronary, coronary artery disease, artery/arterial and angina were found important; for atrial fibrillation flutter, fibrillation, atrial, fibrillation, atrial fibrillation and paroxysm were found important. Meanwhile, the results based on the two metrics were similar, indicating that the importance of words was relatively stable over the thirty bootstrapping experiments. To conclude, the models based on TF-IDF and LR predicted I-codes not only had high AU-ROC and AUPRC, but were also interpretable based on clinically meaningful terms determining the prediction.

**Table 1:**
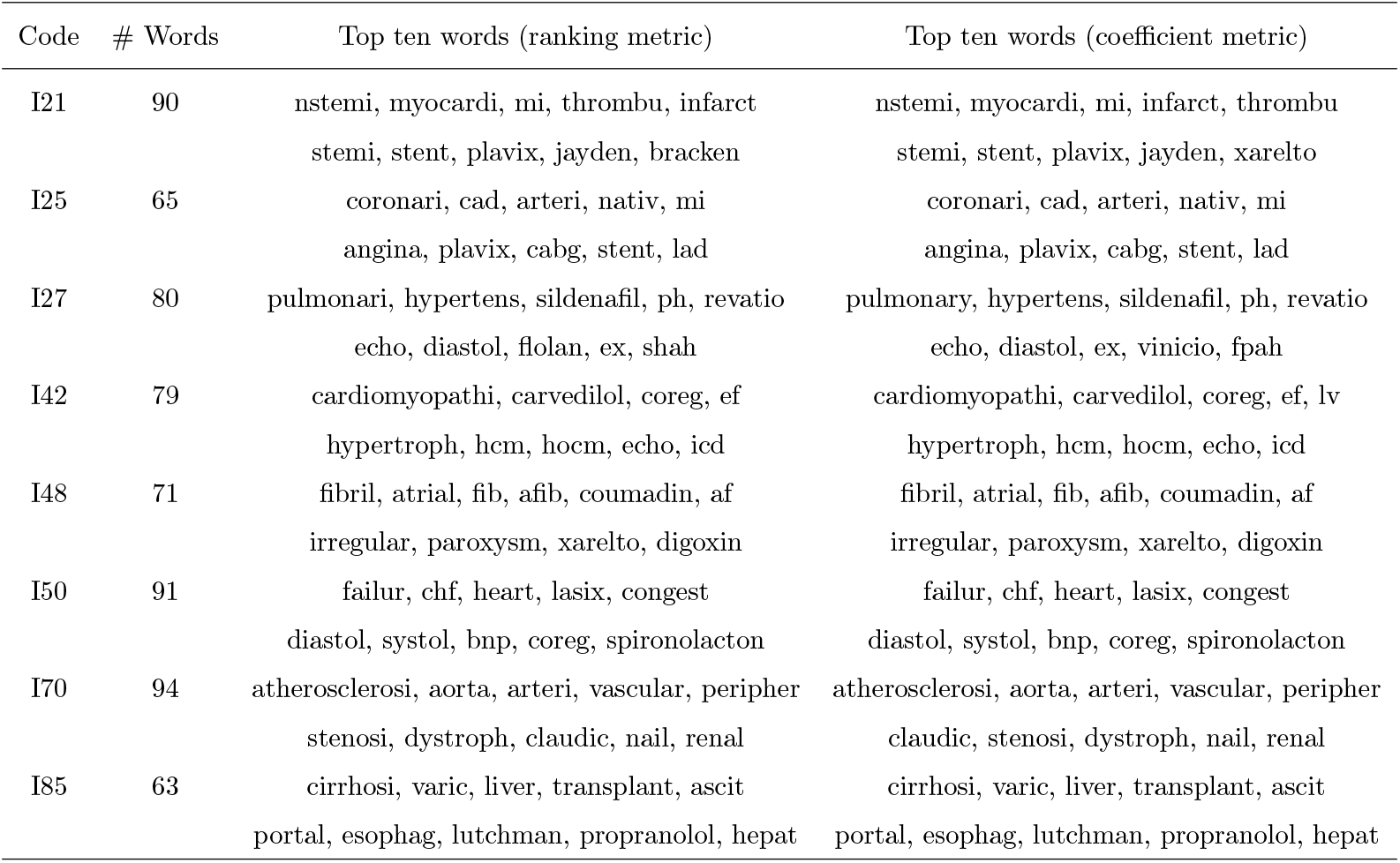
The ten most important words found in thirty bootstrapping experiments based on ranking metric and coefficient metric with TF-IDF and LR. The ranking metric ranks the important words by the sum of the rankings of the word importance in bootstrapping experiments and the coefficient metric ranks the important words by the sum of LR coefficients in bootstrapping experiments. The words are shown after stemming.

### False positive analysis of the prediction

Next, to test whether there were missing diagnostic codes in the data sets that could be imputed by the I-code prediction models, several randomly selected false positive cases were analyzed (Table 2). This analysis suggests that it is possible to impute missing I-codes based on the model predictions in a subset of cases. Additional manual curation efforts might be needed because the most accurate TF-IDF embedding was word-based that has problems dealing with negation, personal and family medical history. For instance, an I-code might be predicted due to a patient’s medical history but not necessarily noted down as the diagnostic code for that specific encounter.

**Table 2:** The analysis of false positive cases based on TF-IDF. The note predictions were manually analyzed and labeled as true (T) or false (F) and the potential causes of erroneous predictions were listed.

| Code | Evidence excerpts of note | T/F | Issue |
| --- | --- | --- | --- |
| I25 | "...all negative for stress induced ischemia..." | F | Negation |
| I48 | "...has the following active medical issues hx of afib..." | T |  |
| I48 | "...paroxysmal atrial fibrillation is seen here..." | T |  |
| I48 | "...was found to be in atrial fibrillation..." | T |  |
| I48 | "...cardiac history of atrial flutter and atrial fibrillation..." | F | Personal Medical History |
| I48 | "...for post hospital check after admission for atrial fibrillation..." | F | Personal Medical History |

### Model transferability on MIMIC-III data set

To test the model transferability, we extracted the discharge summaries in MIMIC-III data set and the corresponding ICD-9 diagnostic codes of each of the eight ICD-10 codes, and tested the pre-trained word embedding models and classification models on the MIMIC-III data set without any fine-tuning. The high AUROC and AUPRC values showed that all models (i.e. TF-IDF, W2V, W2V batch and D2V) models could be well transferred to the classification of the diagnostic codes in the MIMIC-III data set (Fig. 5, Table 3). TF-IDF showed the best performances with the highest AUROC values and AUPRC values while however, BOW performed the worst in terms of AUROC and AUPRC on the majority of the classification tasks. When compared with the test set of Cohort 2, the TF-IDF models reached higher AUPRC values on I21, I25, I48, I50, I85. Generally according to the results, besides BOW models, the other models generalized well to an external cohort.

**Table 3:**
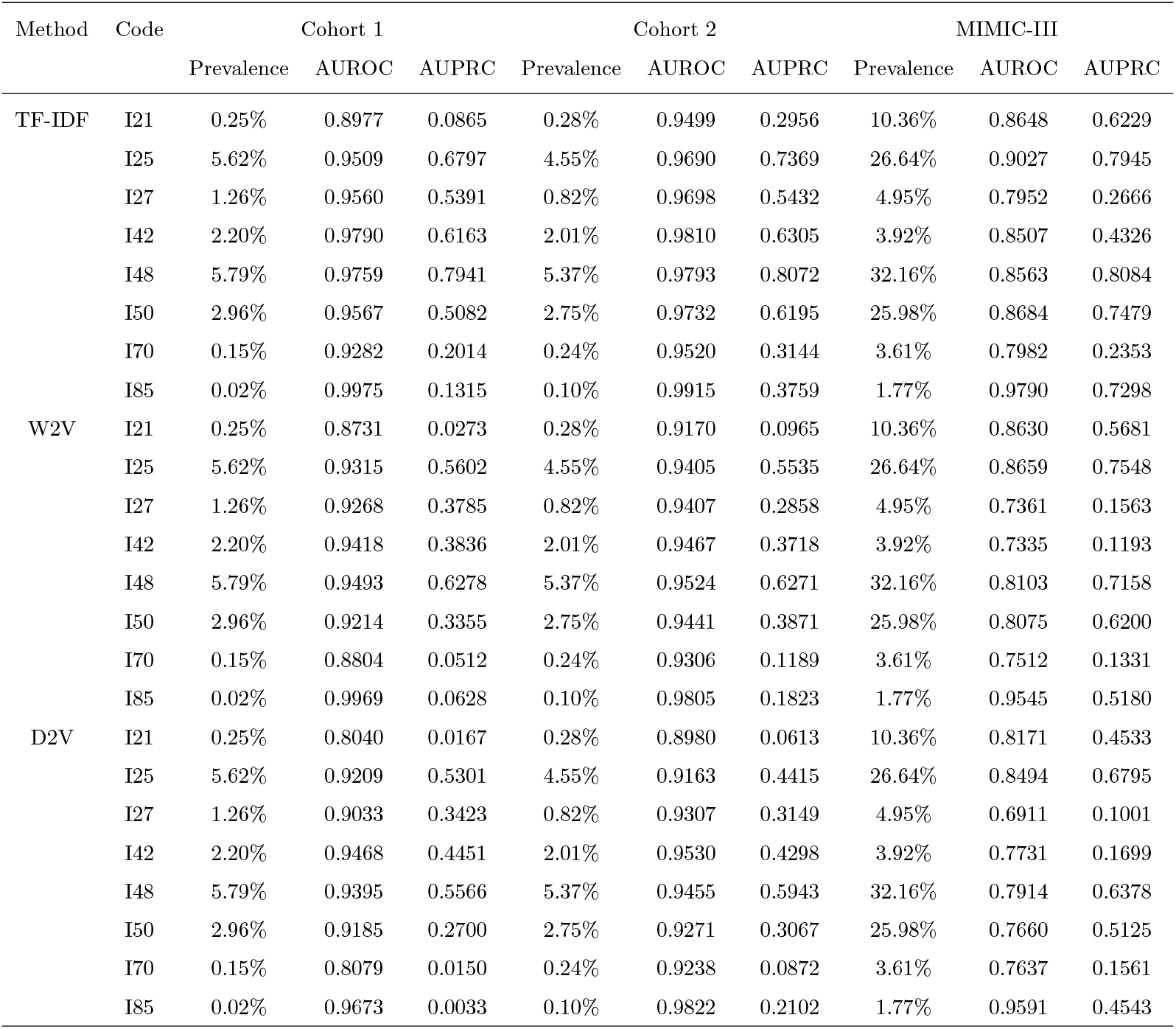
Prevalence and prediction performance on test sets of Stanford cohorts and MIMIC-III data set based on TF-IDF, W2V and D2V.

| Method | Code | Cohort 1 |  |  | Cohort 2 |  |  | MIMIC-III |  |  |
| --- | --- | --- | --- | --- | --- | --- | --- | --- | --- | --- |
|  |  | Prevalence | AUROC | AUPRC | Prevalence | AUROC | AUPRC | Prevalence | AUROC | AUPRC |
| TF-IDF | I21 | 0.25% | 0.8977 | 0.0865 | 0.28% | 0.9499 | 0.2956 | 10.36% | 0.8648 | 0.6229 |
|  | I25 | 5.62% | 0.9509 | 0.6797 | 4.55% | 0.9690 | 0.7369 | 26.64% | 0.9027 | 0.7945 |
|  | I27 | 1.26% | 0.9560 | 0.5391 | 0.82% | 0.9698 | 0.5432 | 4.95% | 0.7952 | 0.2666 |
|  | I42 | 2.20% | 0.9790 | 0.6163 | 2.01% | 0.9810 | 0.6305 | 3.92% | 0.8507 | 0.4326 |
|  | I48 | 5.79% | 0.9759 | 0.7941 | 5.37% | 0.9793 | 0.8072 | 32.16% | 0.8563 | 0.8084 |
|  | I50 | 2.96% | 0.9567 | 0.5082 | 2.75% | 0.9732 | 0.6195 | 25.98% | 0.8684 | 0.7479 |
|  | I70 | 0.15% | 0.9282 | 0.2014 | 0.24% | 0.9520 | 0.3144 | 3.61% | 0.7982 | 0.2353 |
|  | I85 | 0.02% | 0.9975 | 0.1315 | 0.10% | 0.9915 | 0.3759 | 1.77% | 0.9790 | 0.7298 |
| W2V | I21 | 0.25% | 0.8731 | 0.0273 | 0.28% | 0.9170 | 0.0965 | 10.36% | 0.8630 | 0.5681 |
|  | I25 | 5.62% | 0.9315 | 0.5602 | 4.55% | 0.9405 | 0.5535 | 26.64% | 0.8659 | 0.7548 |
|  | I27 | 1.26% | 0.9268 | 0.3785 | 0.82% | 0.9407 | 0.2858 | 4.95% | 0.7361 | 0.1563 |
|  | I42 | 2.20% | 0.9418 | 0.3836 | 2.01% | 0.9467 | 0.3718 | 3.92% | 0.7335 | 0.1193 |
|  | I48 | 5.79% | 0.9493 | 0.6278 | 5.37% | 0.9524 | 0.6271 | 32.16% | 0.8103 | 0.7158 |
|  | I50 | 2.96% | 0.9214 | 0.3355 | 2.75% | 0.9441 | 0.3871 | 25.98% | 0.8075 | 0.6200 |
|  | I70 | 0.15% | 0.8804 | 0.0512 | 0.24% | 0.9306 | 0.1189 | 3.61% | 0.7512 | 0.1331 |
|  | I85 | 0.02% | 0.9969 | 0.0628 | 0.10% | 0.9805 | 0.1823 | 1.77% | 0.9545 | 0.5180 |
| D2V | I21 | 0.25% | 0.8040 | 0.0167 | 0.28% | 0.8980 | 0.0613 | 10.36% | 0.8171 | 0.4533 |
|  | I25 | 5.62% | 0.9209 | 0.5301 | 4.55% | 0.9163 | 0.4415 | 26.64% | 0.8494 | 0.6795 |
|  | I27 | 1.26% | 0.9033 | 0.3423 | 0.82% | 0.9307 | 0.3149 | 4.95% | 0.6911 | 0.1001 |
|  | I42 | 2.20% | 0.9468 | 0.4451 | 2.01% | 0.9530 | 0.4298 | 3.92% | 0.7731 | 0.1699 |
|  | I48 | 5.79% | 0.9395 | 0.5566 | 5.37% | 0.9455 | 0.5943 | 32.16% | 0.7914 | 0.6378 |
|  | I50 | 2.96% | 0.9185 | 0.2700 | 2.75% | 0.9271 | 0.3067 | 25.98% | 0.7660 | 0.5125 |
|  | I70 | 0.15% | 0.8079 | 0.0150 | 0.24% | 0.9238 | 0.0872 | 3.61% | 0.7637 | 0.1561 |
|  | I85 | 0.02% | 0.9673 | 0.0033 | 0.10% | 0.9822 | 0.2102 | 1.77% | 0.9591 | 0.4543 |

**Figure 5:**
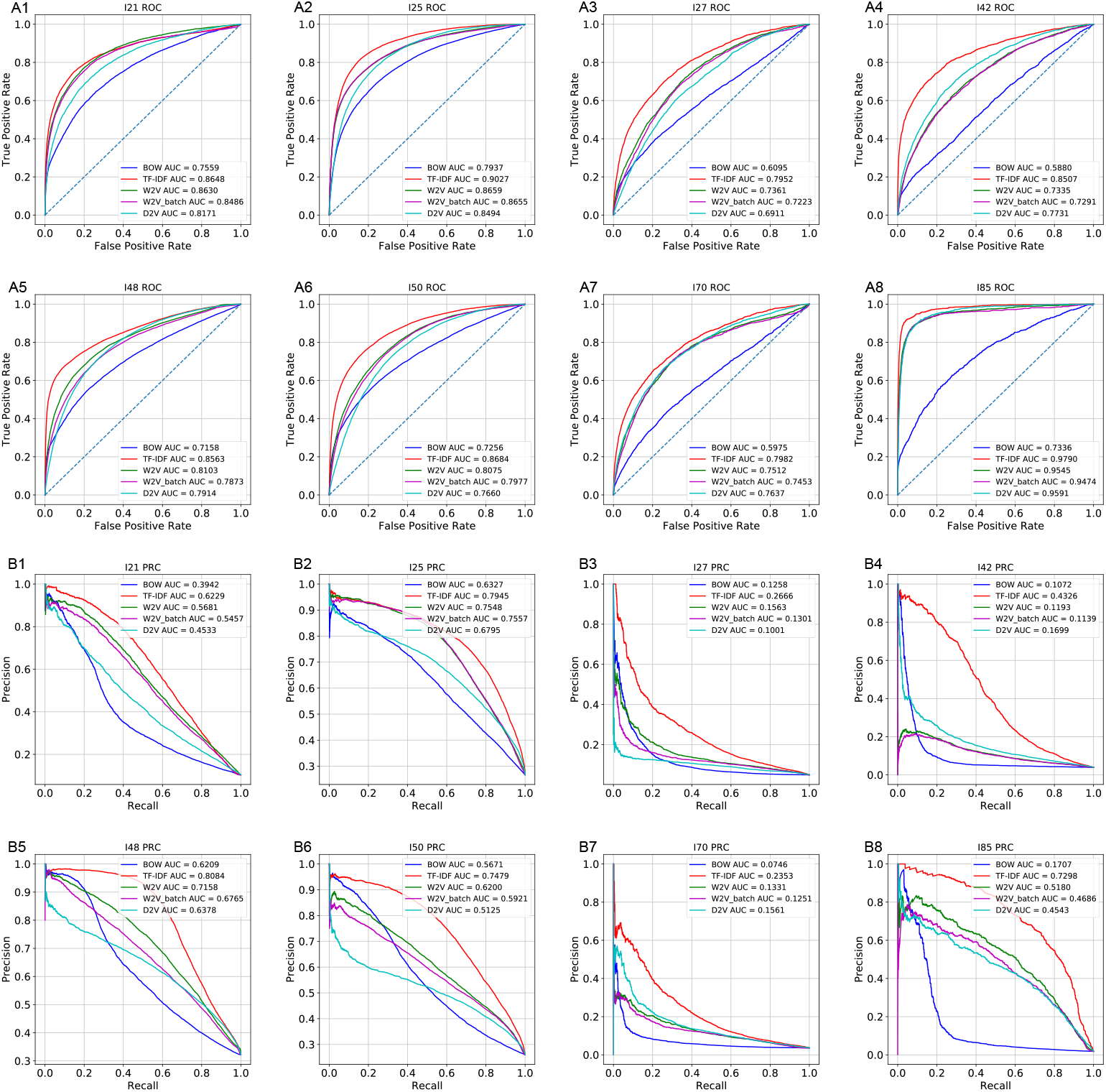
The receiver operating characteristic curves and precision recall curves in the classification of eight corresponding ICD-9 codes on the discharge summary in MIMIC-III data set. A1-A8: the classification AUROC. B1-B8: the classification AUPRC.

## Discussion

In this work, natural language processing (NLP) methods were used to compare five different word embeddings from free-text outpatient clinical notes and then LR was shown effective to predict the diagnostic codes of eight cardiovascular diseases. Among them, on both the smaller Cohort 1 and the larger Cohort 2 from the Stanford EHR data set, the best embedding according to AUROC and AUPRC was TF-IDF (Figs. 3, 4, Supplementary Figs. 1, 2). From the Cohort 1 to the Cohort 2, the scalability of the models was shown that with more data the prediction performance could be improved (Fig. 3, Supplementary Fig. 4). Additionally, the majority of the embedding models and classification models trained on the Stanford EHR data set also showed transferability when applied to the MIMIC-III data set (Table 3). The TF-IDF, W2V, W2V batch and D2V models performed well on the Stanford cohorts and generalized well on the different MIMIC-III data set. The simple BOW embeddings showed a sharp decrease in AUROC and AUPRC values on the different data set, showing that the embedding models might have overfitted the Stanford EHR data set because it directly used the word counts as features without any normalization and the distribution of the word counts in different data sets is likely to be different. The word embedding with normalization on word counts (TF-IDF) and the word em-beddings (W2V, D2V) that seeks for a lower-dimension representation showed higher robustness in classification performance when transferred to a different data set, potentially because the normalization and the reduced dimensionality may lead to smaller variance across data sets. The results imply that these models can be used in accurately predicting diagnostic codes and improving the quality of diagnostic codes at different clinical sites. Furthermore, although the new word embeddings (W2V, W2V batch and D2V) embeddings did not show higher AUROC and AUPRC when compared with TF-IDF, they were in lower dimensions (200/600) than TF-IDF and BOW(414,391), which could be helpful to significantly reduce computational costs with fair classification performance in AUROC and AUPRC.

Additionally, the interpretability of the models was shown in this work with important word analysis and false-positive-case analysis. The important words found in each I-code prediction tasks were clinically meaningful (Table 1). The robustness of the important words was also shown with bootstrapping. In a previous study, Wei et al [1] applied convolutional neural network to predict diagnostic ICD-10 codes with good performances but the deep-learning based models were hard to interpret. Sheikhalishahi et al. [2] also mentioned in their review paper that the model interpretability was a significant issue for more complex methods. Wei et al. [1] claimed that the simple word embedding do not give good results and showed the CNN-embedding with SVM reached a precision value of 0.2162 and a recall value of 0.7732 in the prediction of diagnostic codes. Although direct benchmarking and comparison cannot be made due to differences in the prevalence of ICD-10 codes and data sets selected in this study, the simple word vectorization models and LR showed good predictive performances in our study (Fig. 3, Table 3), while maintaining interpretability and therefore could contribute to the diagnostic code prediction and quality control for clinicians.

Next, false positive case analysis showed that some of the false positive pre-dictions might be correct and could be applied to impute potential missing codes that do not have I-codes recorded by clinicians (Table 2). The false positive pre-dictions might not be wrong but are simply missing. However, among the false positive cases, we also observed that certain mistakes were caused by negation, past medical and family history. Because the best model of TF-IDF method is word-based, it models the contents of the free-text by each individual word, and these issues cannot be directly detected by the TF-IDF model. Therefore, to impute missing I-codes, the proposed classifiers here could be used to complete records, in combination with additional methods to assert negation, temporality and who the experiencer is.

More generally, an important use case of this work is to impute ICD-10 codes from unstructured free-text format. As diagnostic codes rich in clinical information can be missing and the noted diagnostic codes may also be inaccurate, which has been showed by recent studies for diagnostic codes related to my-ocardial infarction and stroke [13, 12]. This study proposes a method to impute missing diagnostic codes and potentially correct misclassified diagnostic codes based on model predictions. In addition, the model interpretability also enables clinicians to interpret the models and check whether particular imputation is correct. The improvement of the quality of diagnostic codes may help further machine learning diagnosis because machine learning algorithms typically require structured data. Many of the previous studies directly use the diagnostic codes for the following downstream classification tasks [1, 2, 9, 10]. To improve the quality of diagnostic codes also could improve the data quality for further machine learning processes.

Although this study has shown promising results of predicting diagnostic codes based on clinical notes, there are several points in this study that could be further studied. Firstly, our modification of segregating the texts into batches did not improve the performances when compared with conventional word2vec model. The probable reason may be the notes are of different lengths and different lengths of sections. Roughly splitting the notes into fixed batches may not successfully partition the different sections. In the future, studies can be designed to automatically detect and partition sections to improve the classification performance. Secondly, in this study, after the first step of data processing, 63.2% notes were removed because they either didn’t have a diagnostic code or were shorter than 60 words. We used these dropped notes in training the doc2vec embedding models. The part of unlabeled notes might still contain meaningful information related to classification. Such methods as semi-supervised learning and conformal predictions [30, 31] might be hold potential to make use of these unlabeled data, which could potentially further improve the prediction performance. Thirdly, this work focused on the prediction of ICD-10 codes and the structured codes was not tested in downstream tasks such as phenotyping or outcome prediction with machine learning. This work might help subsequent prediction tasks. For example, the structured diagnostic codes based on the information from clinical notes, can be combined with other data sources in data fusion tasks including imaging data, genomics data and laboratory test data to predict prognosis, patient outcome and disease subtypes [32, 33, 34, 35].

## Data Availability

The data used in this study is not shareable as the data concerns patient information.

## Acknowledgements

This research used data or services provided by STARR, the STAnford medicine Research data Repository, a clinical data warehouse containing live Epic data from Stanford Health Care (SHC), the University Healthcare Alliance (UHA) and Packard Children’s Health Alliance (PCHA) clinics and other auxiliary data from Hospital applications such as radiology PACS. STARR platform is developed and operated by Stanford Medicine Research IT team and is made possible by Stanford School of Medicine Research Office. Research reported in this publication was partially funded by the National Institute of Biomedical Imaging and Bioengineering of the National Institutes of Health (NIBIB), R01 EB020527, and R56 EB020527. The content is solely the responsibility of the authors and does not necessarily represent the official views of the National Institutes of Health. This work is also supported by the Stanford Department of Bioengineering.

## Author Contributions

X. Z., M. HD., P. M. and O. G. conceived the study. O. G. collected the data. X. Z. did the experiments, analyzed the data and wrote the manuscript. M. HD., P. M. and O. G. supervised the work and revised the manuscript. O. G. provided the funding of this work.

## Data Availability

The data used in this study is not shareable as the data concerns patient information.

