## Supplementary materials for "Structuring clinical text with AI: old vs. new natural language processing techniques evaluated on eight common cardiovascular diseases"

*Bag-of-words (BOW) embedding*

Bag-of-words (BOW) [1] is a word-count based embedding algorithm which is commonly used in document classification. The method counts the frequency of each term in the text and uses the frequency of individual term as the feature. The number of features is the same as the number of all distinct terms in the training set and the feature values are proportional to the occurrences of the distinct terms.

*Term frequency-inverse document frequency (TF-IDF) embedding*

Term frequency-inverse document frequency (TF-IDF) [2] is an algorithm with normalized BOW embeddings to emphasize the different importance of terms. The feature in TF-IDF is the ratio of term frequency (TF) and inverse document frequency (IDF). The value of a term embedding increases proportionally to the term frequency but is offset by the number of texts that contain the term. The feature dimensions were the same as those of BOW.

*Word2vec (W2V) embedding*

Word2vec (W2V) [3] is a vectorization algorithm to get word embeddings. Instead of directly using the terms as features, W2V learns an embedding matrix E that contains the embedding of all the terms appearing in the training corpus and maps each term to a feature vector. The embedding matrix, as a goal of optimization, is learned through shallow, two-layer neural networks on simple prediction tasks. Continuous bag-of-words (CBOW) and continuous skip-gram are the two models that can be used to learn the embedding matrix. In the CBOW, the task of the neural network is to predict the current term based on its neighboring terms. In continuous skip-gram, the task is to predict the surrounding terms based on the current term.

*t-distributed stochastic neighbor embedding (tSNE)*

t-distributed stochastic neighbor embedding (tSNE) [4] is a nonlinear dimensionality reduction method which could embed high-dimensional data into 2D space for data visualization by minimizing the Kullback-Leibler divergence (KL divergence) between the low-dimensional distribution and the high-dimensional distribution. In this study, considering the high feature dimensionality, principal component analysis (PCA) was used to firstly lower the dimension to 100 before t-SNE was applied for faster calculation.

*Supplementary Tables and Figures*

1) The prevalence of eight common cardiovascular diseases and ICD-10 codes in the training set, validation set and test set in Cohort 1 and Cohort 2 was shown in Supplementary Table 1.

2) The receiver operating characteristic curves and precision recall curves on eight I-code prediction tasks on Cohort 1 were shown in Supplementary Fig. 1.

3) The receiver operating characteristic curves and precision recall curves on eight I-code prediction tasks on Cohort 2 were shown in Supplementary Fig. 2.

4) The example hyperparameter tuning results were shown in Supplementary Fig. 3, with each subplot showing the receiver operating characteristics curves of TF-IDF LR classifier with varying strengths of L2 regularization.

5) The improvement in AUROC and AURPC of the TF-IDF word embedding and word2vec word embedding from Cohort 1 to Cohort 2 was shown in Supplementary Fig. 4.

6) The example confusion matrices based on LR and TF-IDF embedding (Cohort 1) were shown in Supplementary Fig. 5.

7) The distribution of different categories of encounter and the histogram of note length in the cardiovascular outpatient progress notes data set of Stanford EHR were shown in Supplementary Fig. 6.

Supplementary Table 1: The prevalence of eight common cardiovascular diseases and ICD-10 codes in Cohort 1 and Cohort 2.

| Cohort | Code | Description | Training | Validation | Test |
|---|---|---|---|---|---|
| 1 | I21 | Acute myocardial infarction | 0.26% | 0.24% | 0.25% |
| | I25 | Chronic ischemic heart disease | 4.46% | 4.74% | 5.62% |
| | I27 | Other pulmonary heart disease | 0.82% | 1.03% | 1.26% |
| | I42 | Cardiomyopathy | 1.90% | 1.80% | 2.20% |
| | I48 | Atrial fibrillation flutter | 5.76% | 5.23% | 5.79% |
| | I50 | Heart failure | 2.97% | 3.26% | 2.96% |
| | I70 | Atherosclerosis | 0.29% | 0.26% | 0.15% |
| | I85 | Esophageal Varices | 0.12% | 0.12% | 0.02% |
| 2 | I21 | Acute myocardial infarction | 0.25% | 0.26% | 0.28% |
| | I25 | Chronic ischemic heart disease | 4.69% | 4.45% | 4.55% |
| | I27 | Other pulmonary heart disease | 0.84% | 0.89% | 0.82% |
| | I42 | Cardiomyopathy | 1.87% | 1.79% | 2.01% |
| | I48 | Atrial fibrillation flutter | 5.16% | 5.02% | 5.37% |
| | I50 | Heart failure | 2.70% | 2.55% | 2.75% |
| | I70 | Atherosclerosis | 0.25% | 0.28% | 0.24% |
| | I85 | Esophageal Varices | 0.12% | 0.09% | 0.10% |

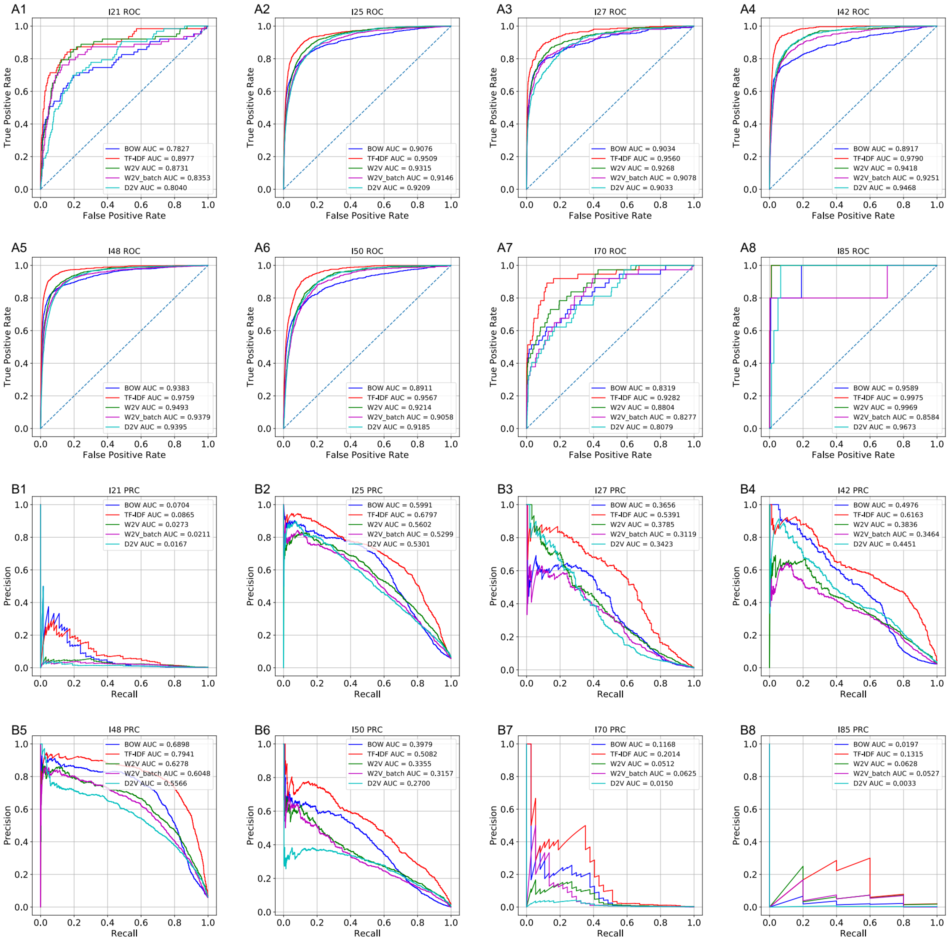


Supplementary Figure 1: The receiver operating characteristic curves and the precision recall curves of the LR models trained on different word embeddings and on the eight I-code classification tasks (Cohort 1).

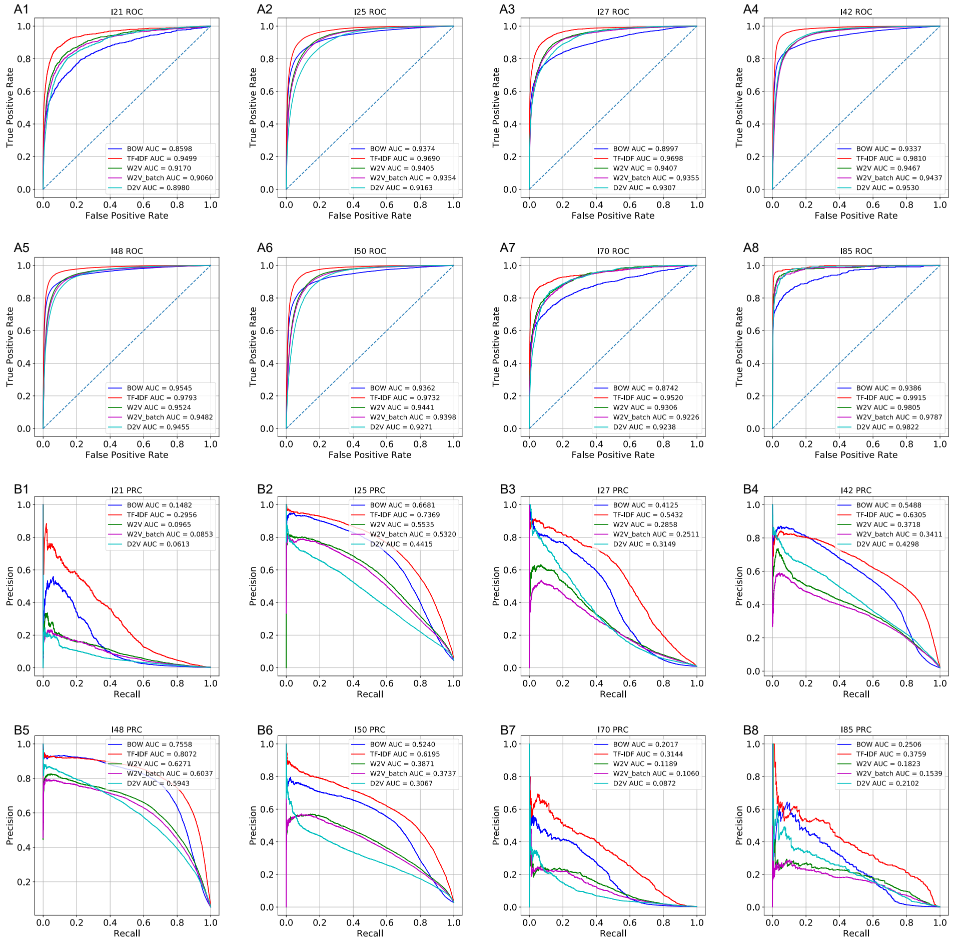

Supplementary Figure 2: The receiver operating characteristic curves and the precision recall curves of the LR models trained on different word embeddings and on the eight I-code classification tasks (Cohort 2).

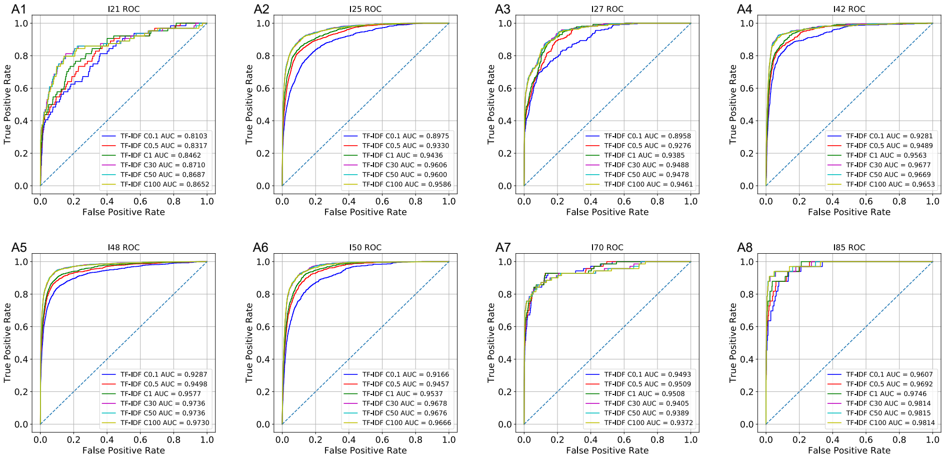


Supplementary Figure 3: The receiver operating characteristic curves of TF-IDF word embedding and Logistic Regression classifier with varying strength of L2 regularization. $C$ was the coefficient that denoted the inverse of the strengths of penalty. Six different $C$ values were tested on the validation set of Cohort 1 and the maximum average AUROC over eight classification tasks was given by $C = 30$

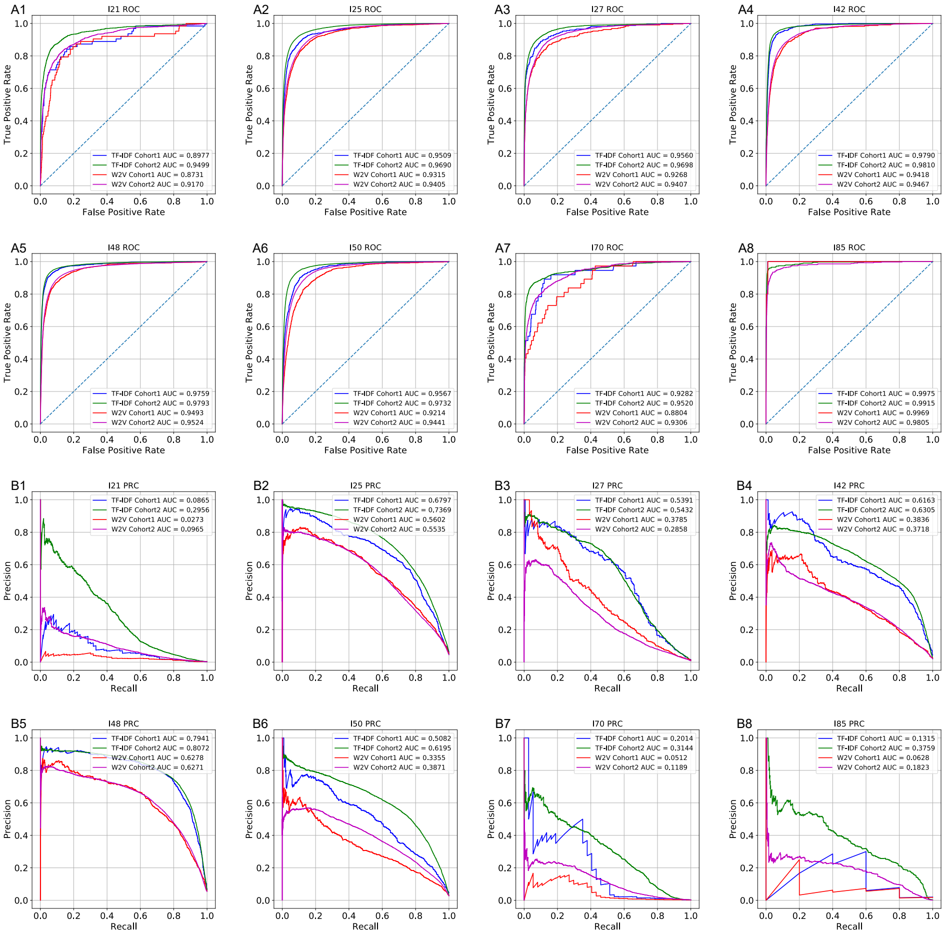


Supplementary Figure 4: The receiver operating characteristic curves and the precision recall curves of the LR models trained on TF-IDF and W2V word embeddings on the eight I-code classification tasks (Cohort 1 and Cohort 2).

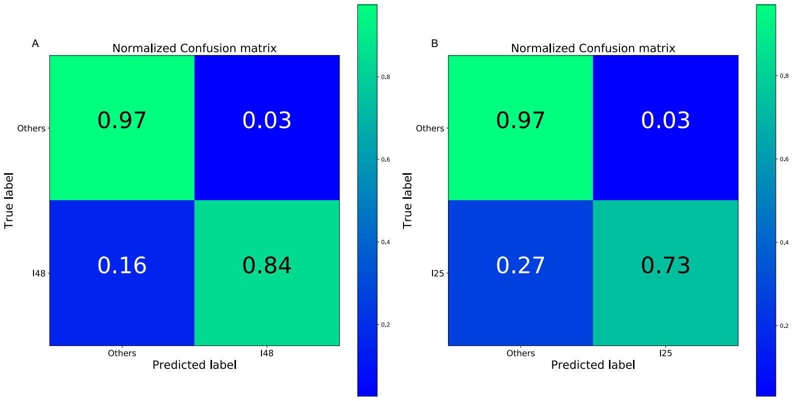


Supplementary Figure 5: The example confusion matrices based on LR and TF-IDF embedding (Cohort 1). The classification thresholds were chosen with a grid search by setting a threshold on sensitivity (0.8 for I48 and 0.7 for I25) and optimizing precision. A. The confusion matrix for I48 prediction. The sensitivity was 0.8401 and the precision was 0.6654. B. The confusion matrix for I25 prediction. The sensitivity was 0.7338 and the precision was 0.5907

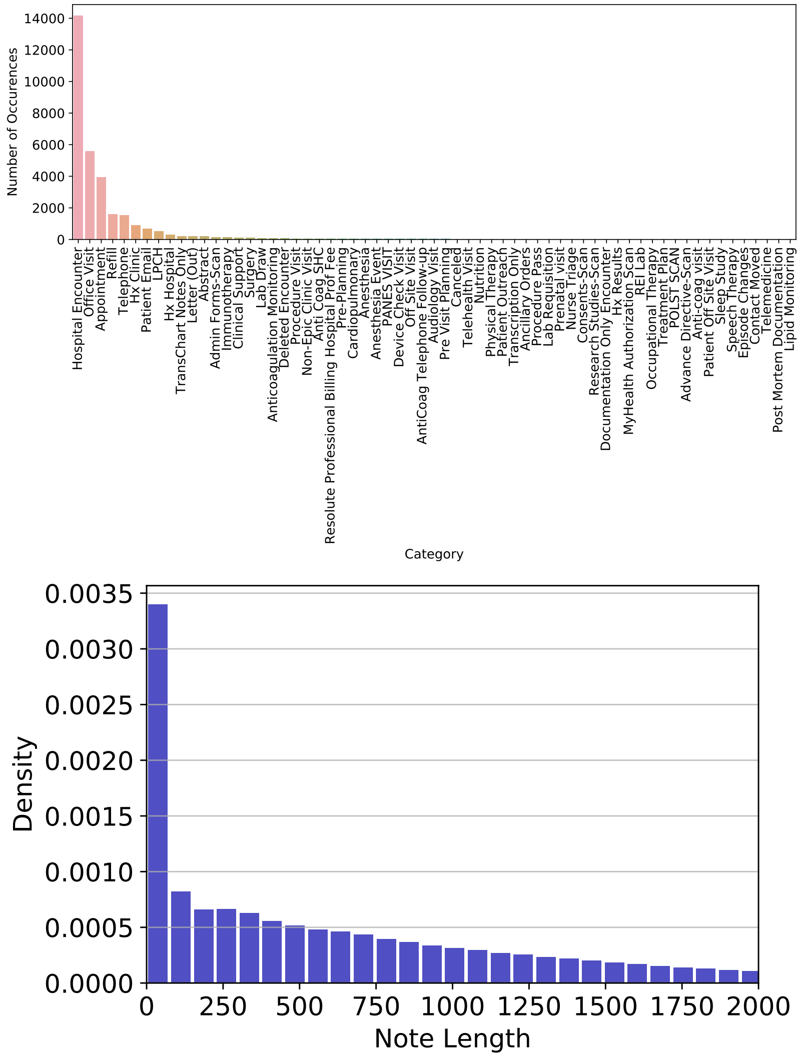


Supplementary Figure 6: The distribution of different categories of encounter and the histogram of note length in the cardiovascular outpatient progress notes data set of Stanford EHR.

[2] K. S. Jones, A statistical interpretation of term specificity and its application in retrieval, Journal of documentation.

[3] T. Mikolov, I. Sutskever, K. Chen, G. S. Corrado, J. Dean, Distributed representations of words and phrases and their compositionality, in: Advances in neural information processing systems, 2013, pp. 3111–3119.

[4] L. v. d. Maaten, G. Hinton, Visualizing data using t-sne, Journal of machine learning research 9 (Nov) (2008) 2579–2605.
